# Exposome contribution to the brain metabolome: importance of body brain connection

**DOI:** 10.64898/2026.05.05.26352469

**Authors:** Naama Karu, Haoqi Nina Zhao, Richa Batra, Matthias Arnold, Jan Krumsiek, Lurian Caetano David, Dinesh Barupal, Leyla Schimmel, Alexandra Kueider-Paisley, Colette Blach, Kamil Borkowski, Pieter Dorrestein, David A Bennett, Rima Kaddurah-Daouk, the Alzheimer’s Disease Metabolomics Consortium

## Abstract

**INTRODUCTION:** Mounting evidence support exposome influences on brain function and health, complementing genome influences. Understanding the molecular imprint of exposome on brain metabolism and the biochemical communication between the body and brain can impact our fundamental understanding and treatment of neuropsychiatric diseases.

**METHODS:** Leveraging two complementary metabolomics platforms, we classified 1400 features in 514 brains from the ROSMAP collection. We evaluated the origin of these compounds using literature and databases. We correlated those metabolites with cognitive function using linear models.

**RESULTS:** We identified over 230 non-endogenous compounds in the brain, including 103 drugs and metabolites, 120 dietary and microbial products and possibly 15 compounds from environmental exposures. Over 20 dietary and gut microbial compounds showed associations with cognition.

**DISCUSSION:** Comprehensive profiling of chemicals in the brain and the link to cognitive function provides foundational work to connect body and brain in the study of AD and related dementias.

## 1. INTRODUCTION

A growing body of evidence supports exposome contribution to brain health and development of neurodegenerative and neuropsychiatric diseases, complementing genetic influences. The exposome includes environmental pollutants, gut microbiome metabolites, diet, drugs, and socioeconomic factors [1]. The molecular basis for how these exposomic factors influence brain chemical composition and metabolic state remains unknown.

Environmental chemical exposure has become a refocused area in the study of central nervous system (CNS) diseases, including Alzheimer’s disease (AD) [2], with large initiatives recently launched by NIA including TOXAD [3]. Chemical exposures include bisphenols, phthalates, persistent organic pollutants (PCBs, PFAS), heavy metals [4–6] household chemicals [7] and pesticides/herbicides; many of which seem to significantly increase AD risk [8]. Disruption of lithium homeostasis in the brain has been implicated in early AD pathogenesis events [9], leading to a pilot clinical trial that reported lithium as benefitting cognition [10]. Many lipophilic xenobiotics can cross the blood brain barrier (BBB) via passive diffusion, and some polar xenobiotics interact with specialized transporters [11]. Additionally, some are neurotoxic, but their mechanisms of action vary, including BBB disruption, competing with essential metals, altering protein signalling, or inducing amyloidosis [8].

Diet and lifestyle, are big influences on overall health, including brain health. Recently, dietary interventions have emerged as a way to mitigate metabolic risk factors and improve cognitive health. Mediterranean-DASH (MIND) dietary intervention [12, 13] is associated with better cognition [14, 15], slower cognitive decline [16], and lower AD risk [13]. Application of ketogenic diet showed improved memory in AD patients [17] and adjusts metabolic profiles that predispose to cognitive decline [18, 19]. Similarly, clinical trials with mild cognitive impairment (MCI) participants showed that dietary (utilizing Dean Ornish and MIND diets) and lifestyle intervention improves cognitive performance [20, 21]. The gut microbiome and the gut bacterial-human co-metabolism contribute profoundly to brain health through the gut-brain axis, which is not fully characterised [22]. GI inflammation occurs in neurodegenerative diseases such as Parkinson’s disease and AD [23, 24]. Human gut bacterial co-metabolism produces many chemicals that influence brain function, including secondary bile acids (BAs), aromatic amino acid metabolites, branch chain amino acids, short-chain fatty acids, TMAO, and neurotransmitters like histamine, GABA, serotonin, and dopamine [25, 26]. We have shown that secondary BAs are altered in MCI and AD and linked to cognitive decline and brain imaging changes [27–30]. Furthermore, we showed that bacterial indoles correlate strongly with anxiety and brain NMR changes [31]. This building evidence indicates the importance of exposome in brain function and highlights a need to define the molecular basis of how these exposomic factors influence the brain and the extent of their presence in the brain.

Metabolomics provides powerful tools to map dysregulation in metabolic processes related to diseases, including AD [32, 33]. It measures a broad spectrum of xenobiotics, including gut microbiome derivatives, environmental contaminants, and food derivatives [2]. The AD Metabolomics Consortium has pioneered the metabolomic profiling of large, well-characterized AD cohorts, including the AD Neuroimaging Initiative (ADNI), the Religious Orders Study/Memory and Aging Project (ROSMAP), and several animal models for AD, providing a comprehensive picture of central and peripheral metabolic alteration across AD trajectories[34]. We have developed an initial brain metabolome atlas for AD and affected processes in dorsolateral prefrontal cortex (DLPFC) [34]. In a complimentary study we highlighted major brain metabolome differences in two brain regions and differences between two diseases: AD and progressive supranuclear palsy [35].

In this manuscript, we address the potential influence of the exposome on the brain metabolome in the DLPFC region, suggesting how these exposome-related chemicals can end up in the brain and their potential influence on cognitive function. This line of investigation is part of a foundational effort to connect the body and the brain, and rethink AD pathogenesis and therapeutic approaches in which the peripheral and central connect.

## 2. METHODS

### 2.1. Cohort

The Religious Orders Study (ROS) and the Rush Memory and Aging Project (MAP) are two ongoing longitudinal clinicopathologic cohorts established by the Rush Alzheimer’s Disease Center. ROS was initiated in 1994 with participants recruited from religious communities across the United States. MAP began in 1997 and enrolled individuals from diverse educational and socioeconomic backgrounds in northeastern Illinois. Both studies were approved by the Institutional Review Board of Rush University Medical Center. Participants were older adults who consented to annual clinical evaluations and postmortem brain donation. Written informed consent included an Anatomic Gift Act and repository agreement that permitted data and biospecimen sharing. Following enrolment, participants underwent yearly assessments of physical and cognitive function. Detailed metadata is described previously [36] and includes (but is not limited to) age at death, sex, body mass index, medication history (at class level only), APOE genotype, education, cognitive scores, rates of cognitive decline, and clinical diagnosis at death. Neuropathologic evaluations were performed after death, and the postmortem interval recorded. For this study, dorsolateral prefrontal cortex (DLPFC) tissue from 514 participants was used for metabolomic profiling, removed, and processed as described in the **Supplementary Methods**. The samples were kept at -80°C until distribution to the analytical laboratories.

### 2.2. Metabolomics profiling and data processing

#### 2.2.1. Metabolon platform

Sample extraction and instrumental analysis are detailed in the **Supplementary Methods**. Briefly, upon receipt, samples were immediately stored at -80°C until processing that involved protein removal and aliquoting to several fractions (for analysis by various ultra-performance liquid chromatography (UPLC)-MS/MS methods) and evaporating of organic solvent. All methods utilized a Waters ACQUITY UPLC and a Thermo Scientific Q-Exactive high resolution/accurate mass spectrometer interfaced with a heated electrospray ionization (HESI-II) source and Orbitrap mass analyzer, operated either in acidic positive ion conditions or basic negative ion conditions, each on a separate dedicated C18 column. Additionally, one aliquot was analysed via negative ionization following elution from a HILIC column. The MS analysis alternated between MS and data-dependent MS^n^ scans using dynamic exclusion. The scan range varied between methods but covered 70-1000 m/z. Raw data was extracted, peak-identified and QC processed using Metabolon’s hardware and software. Peaks were quantified using area-under-the-curve. For studies spanning multiple days, a data normalization step was performed to correct variation that resulted from instrument inter-day tuning differences. Compounds were identified by comparison to library entries of purified standards or recurrent unknown entities, incorporating retention time/index (RI), mass to charge ratio (*m/z)*, and chromatographic data (including MS/MS spectral data). Initial statistical analysis was conducted as comprehensively described in our earlier publication on the same cohort [34]. A follow-up analysis of a re-curated Metabolon dataset utilising metabolites with more than 20% missingness is described in the **Supplementary Methods**.

#### 2.2.2. UCSD untargeted platform

Sample extraction, instrumental analysis, and data processing and annotation are detailed in the **Supplementary Methods**. Briefly, brain samples were plated onto wooden swabs and stored at −80°C. Extraction was conducted via mechanical homogenization with 50/50 water/methanol (v/v) using a TissueLyser. Dried extracts were sealed and stored at -80°C until LC-MS analysis using a C18 column in a Vanquish UHPLC coupled to a Q Exactive quadrupole-orbitrap mass spectrometer (Thermo Fisher Scientific) with positive heated electrospray ionization. Full scan MS1 spectra were acquired from *m/z* 100-1500 at 35,000 resolution (at *m/z* 200). Data-dependent MS/MS was performed on up to five precursors per

MS1 scan using a 3 *m/z* isolation window, 0.5 *m/z* isolation offset, stepped normalized collision energies (20/30/40%), and 35,000 resolution. Raw files were converted to mzML format using MSConvert (ProteoWizard) and processed in MZmine 2 utilising a strict set of peak detection thresholds and feature grouping, and employing strict blank subtraction to minimise the curation of cross-contamination peaks. Feature annotations were assigned using the GNPS spectral library[37]. To contextualize metabolite origins, spectra were queried against curated reference datasets containing >60,000 microbial monocultures, ∼3,500 food extracts, and ∼500 personal-care products using the Fast Search API within the microbeMASST framework [38, 39], as well as the curated GNPS Drug Library [40]. To evaluate whether brain-detected metabolites also appear in other human tissues, we conducted a spectral comparison across all public untargeted metabolomics datasets in GNPS/MassIVE[41], MetaboLights, and the Metabolomics Workbench.

## 3. RESULTS

### 3.1. Metabolomics approaches

The chemical diversity of the brain was demonstrated by two complementary metabolomics approaches. Classification of compound origin were built on the information delivered by each platform and augmented by searches in leading chemical databases and literature, as indicated in **Tables S1-S4**.

#### 3.1.1. Metabolon platform

The Metabolon profiling of the 514 DLPFC brain samples yielded a total of 1135 peaks in at least one brain sample, of which 78 were not characterised and four were partially characterised, leaving 1053 characterised compounds, some with putative annotation (**Tables S1-S4**). Almost 18% of the fully characterised compounds originated only in non-endogenous sources (**Figure 1**). These pharmaceutical drugs, chemicals, dietary compounds, and microbial metabolites are discussed in detail in the following sections.

**Figure 1.**
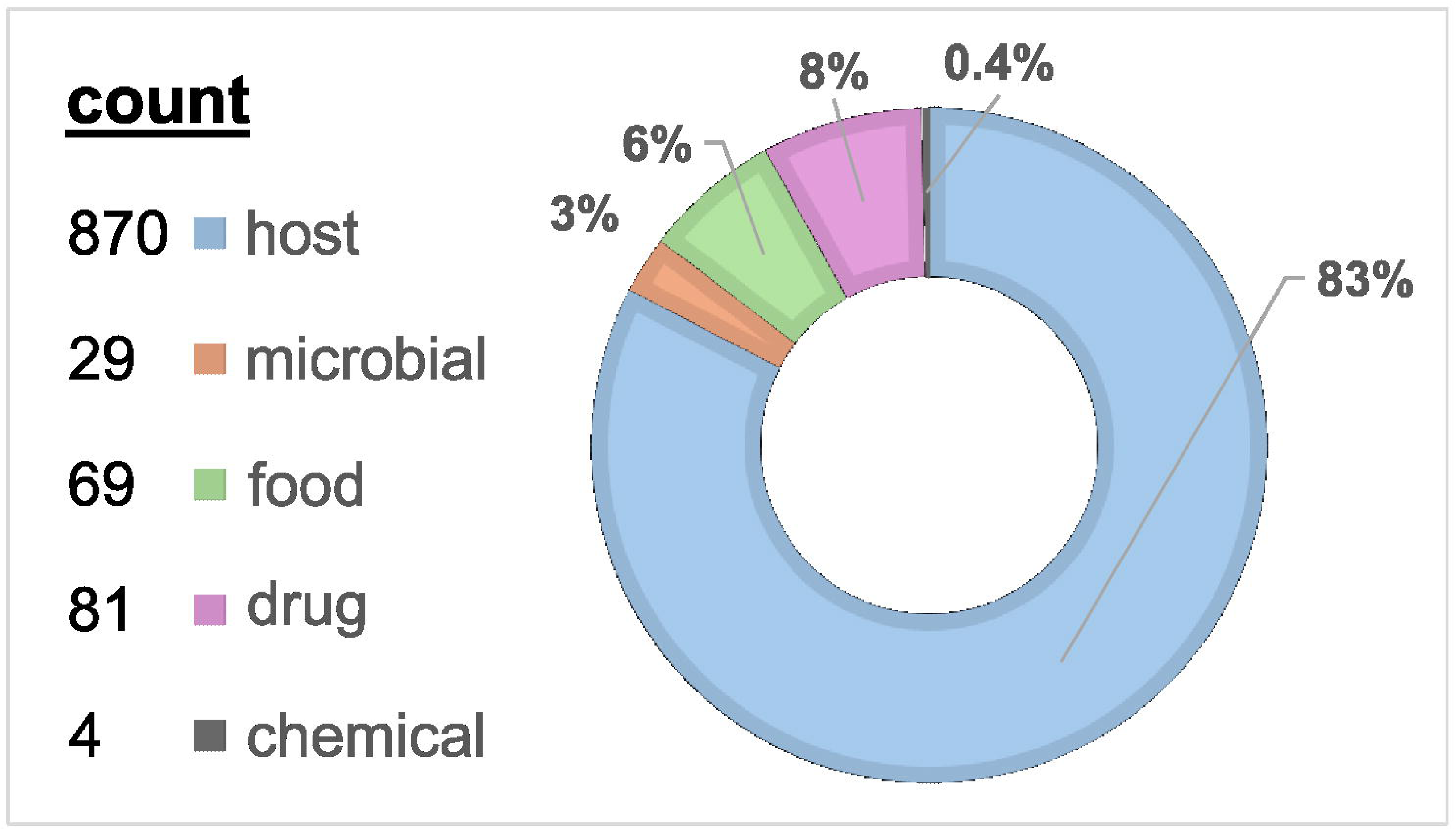
Distribution of compound sources (origin) detected by the Metabolon platform. In case of duplicate origins, the graph shows the most well-established, prominent, or relevant to brain tissue. If a compound is a known endogenous metabolite, it is classified as “host” even if microbes and other organisms can synthesise it, or if it is a dietary component. A diet-microbial-host co-metabolite is mostly presented as “microbial”. Vitamins and their metabolites, as well as essential amino acids and fatty acids, are classified as “diet”. A “chemical” is a compound with an industrial function, or possibly an environmental contaminant.

#### 3.1.2. UCSD untargeted platform

We performed untargeted metabolomics to comprehensively inventory small molecules present in ROSMAP brain samples. In total, 5,653 chemical features with associated MS/MS spectra were detected after blank removal. By matching these spectra against the GNPS community spectral library (annotation level 2/3 following the 2007 Metabolomics Standards Initiative), approximately 7.3% of features were annotated (**Figure 2a**). Among the annotated compounds, 57% were endogenous or microbially derived metabolites (e.g., *N*-acyl lipids, dipeptides, tripeptides), while 31% were exposure-related chemicals. The latter included food-derived compounds (11%; e.g., syringic acid, lenticin, rosmarinic acid), pharmaceuticals and their metabolites (13%; e.g., donepezil, an Alzheimer’s medication; quetiapine, an antipsychotic), and other synthetic chemicals (7%) such as plasticizers (e.g., bis(2-ethylhexyl) phthalate), disinfectants (benzyltetradecyldimethylammonium), and personal care product (PCP) ingredients (e.g., cocamidopropyl betaine).

**Figure 2.**
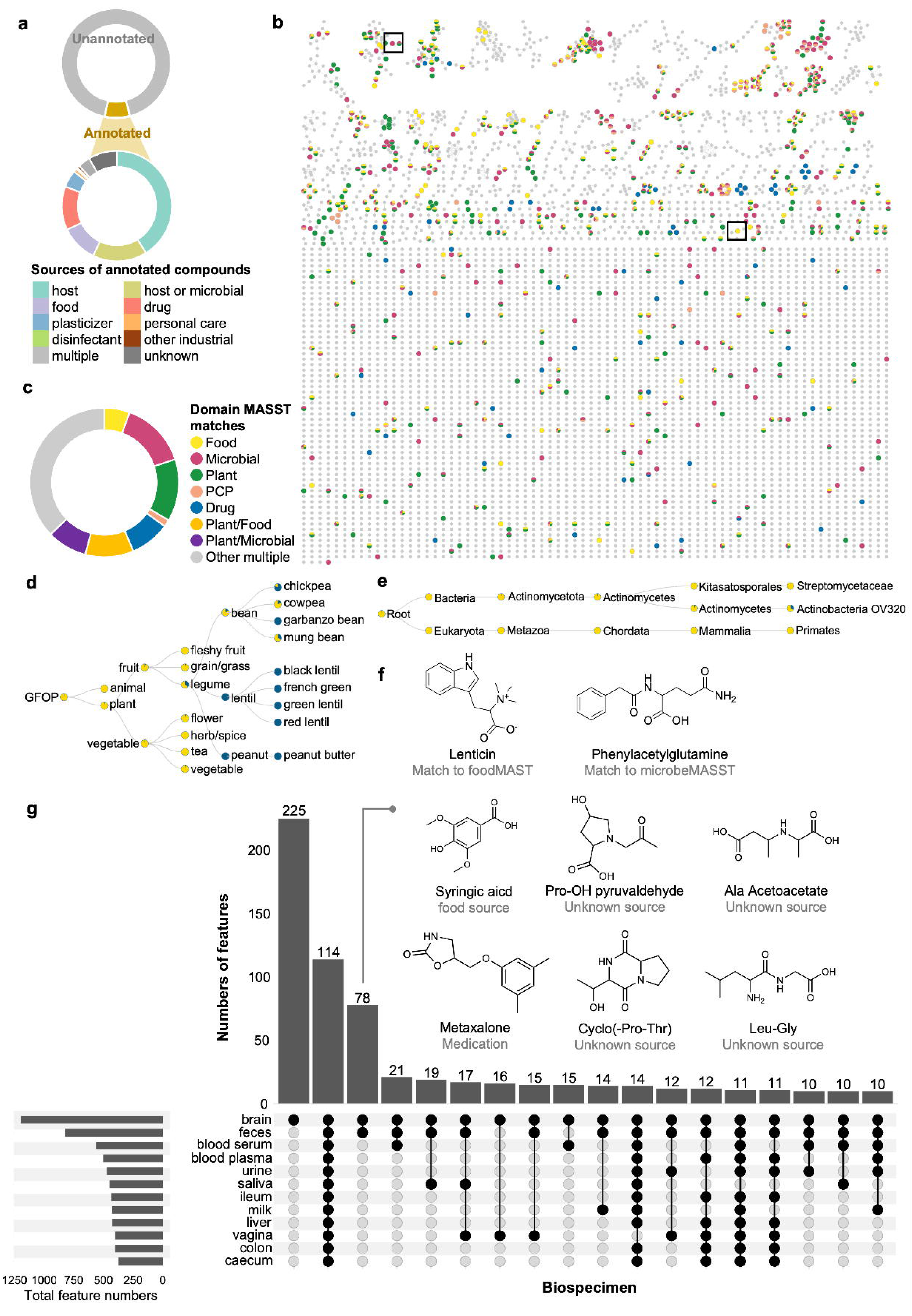
Characterization of molecules in ROSMAP brains through untargeted metabolomics. **a,** Overview of annotated chemical features categorized by potential sources. MS/MS spectra of detected features were searched against the GNPS community spectral library, and annotations were manually classified based on their likely origins. **b,** Molecular network of the detected features. Nodes are colored according to matches with reference datasets: food extracts (yellow), microbial monocultures (purple), plant extracts (green), personal care products (pink), drugs (blue), or no match (gray). **c,** Summary of the number of features matched to each reference dataset. **d,** Lenticin matched to food reference datasets. **e,** Phenylacetylglutamine matched to microbial monocultures. **f,** Chemical structures of lenticin and phenylacetylglutamine . **g,** UpSet plot illustrating the distribution of ROSMAP brain chemical features across other body sites and biospecimen based on integration with ReDU-annotated public datasets, highlighting a subset of molecules shared between brain and feces. *Abbreviations:* GNPS, Global Natural Products Social Molecular Networking; ROSMAP, Religious Orders Study/Memory and Aging Project.

To further infer the potential sources of the detected molecules, including the unannotated ones, we compared the MS/MS spectra from ROSMAP brain samples against a collection of public LC-MS/MS reference datasets that encompassed microbial monocultures, food extracts, plant extracts, PCP formulations, and pharmaceutical compounds (**Figure 2b**). This spectral matching identified 706 putative compound links. Of these, 14% matched exclusively to microbial reference datasets, 5.5% to food references, 8.6% to drugs, and 1.5% to PCPs (**Figure 2c**). The reference matches were consistent with GNPS spectral library annotations. For example, lenticin was linked to food reference spectra, particularly from legume products such as beans, lentils, and peanut products. Phenylacetylglutamine, a gut microbiota-derived metabolite formed by the conjugation of phenylacetate with glutamine, was matched to microbial reference data of Actinomycetes, which are known producers of phenylacetic acid from phenylalanine (**Figure 2d–f**).

To explore molecular cross-talk between the brain and other body sites, we searched all detected chemicals against public metabolomics datasets annotated within ReDU [41], a controlled-vocabulary platform for file-level metadata integration. The most prevalent categories included metabolites detected exclusively in brain tissue or broadly across multiple body parts; notably, the third largest category comprised metabolites shared between brain and feces. Annotated examples include alanine acetoacetate and hydroxyproline pyruvaldehyde - two recently synthesized metabolites from the Dorrestein Lab now reported for the first time in human tissues [42]; as well as syringic acid (a food-derived metabolite), metaxalone (a muscle relaxant), and several dipeptides formed through microbial protease activity (**Figure 2g**). Collectively, these results reveal a widespread presence of diet-derived, microbial, and medication-related molecules in the human brain.

### 3.2. Contribution of drugs to the brain metabolome

Together, the UCSD and Metabolon platforms identified 103 drugs and related metabolites in the brain samples. The UCSD untargeted platform annotated 102 drug-related chemicals in brain samples; however, 54 of them could also originate in endogenous metabolites, diet, or PCPs, and thus were excluded from further analysis. Of the remaining 48 annotated drugs, 25 overlapped with the 80 drugs detected by the Metabolon platform, which suggests a level of orthogonality between the two methods (see **Table S2**). The class distribution of the drugs detected in the brain (**Figure 3**) reflects the expected dominance of drugs that target the CNS and are designed to cross the BBB.

**Figure 3.**
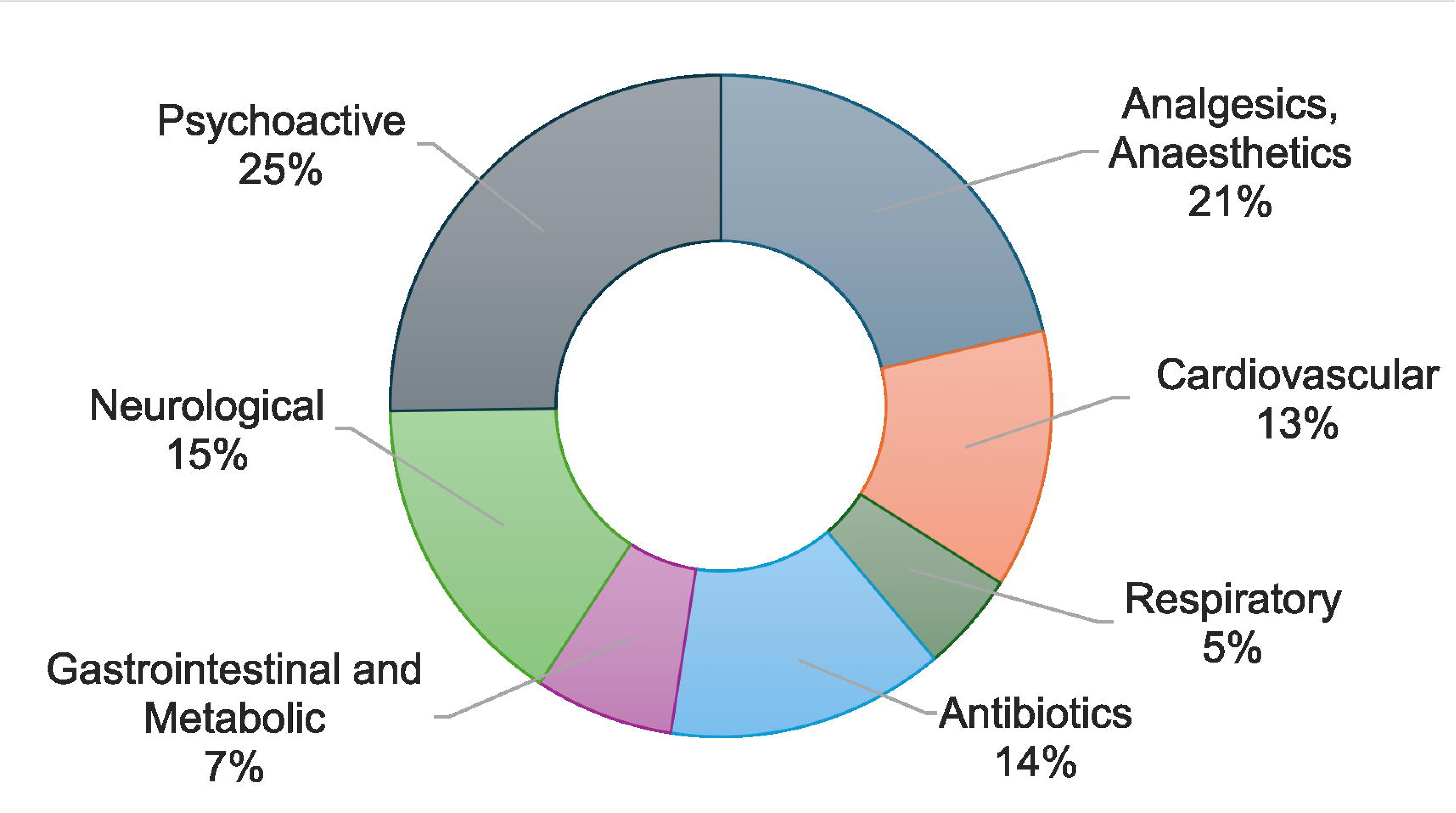
Class distribution of pharmaceutical drugs detected in ROSMAP brains. The testing detected 103 annotated drugs and metabolites. metabolic drugs include anti-gout and anti-diabetic medications. Note that the distribution is by class only and does not reflect the frequency of detection in samples.

The more frequently detected psycho-neurological drugs included the AD medications memantine and donepezil, anticonvulsant gabapentin, the tetracyclic antidepressant mirtazapine, and the SSRI antidepressants citalopram (with 4 metabolites) and sertraline (with one metabolite). The drug class detected in the highest number of samples was analgesics and anaesthetics, possibly reflecting palliative comfort care. All of its 22 compounds were detected by Metabolon, originated in seven drugs, and led by paracetamol (in 414 samples) with eight metabolites, tramadol, lidocaine, and their respective metabolites. Other common drugs included the cardiovascular medication metoprolol and its metabolite (n>100), the antibiotic ofloxacin, and the diuretic furosemide (∼50 each). Participants showed distinct medication exposure profiles based on the normalized LC-MS peak areas – some individuals were exclusively exposed to one drug category (especially neurology and cardiology drugs), while other participants were exposed to multiple drug classes. Moreover, the computationally derived drug metabolite library of UCSD enabled the additional annotation of 15 drug metabolites with high confidence, enhancing the accuracy of medication read-out in untargeted metabolomics. For example, a de-ethylation metabolite of chloroquine and a N-dealkylation metabolite of hydroxyzine were observed in several samples, in the absence of their corresponding precursors.

### 3.3. Contribution of diet and gut microbiota to the brain metabolome

The brain metabolome profile obtained by both platforms together covered approximately 120 metabolites derived from diet and microbial metabolism, as summarised in **Figure 4** and detailed in **Table S3**.

**Figure 4.**
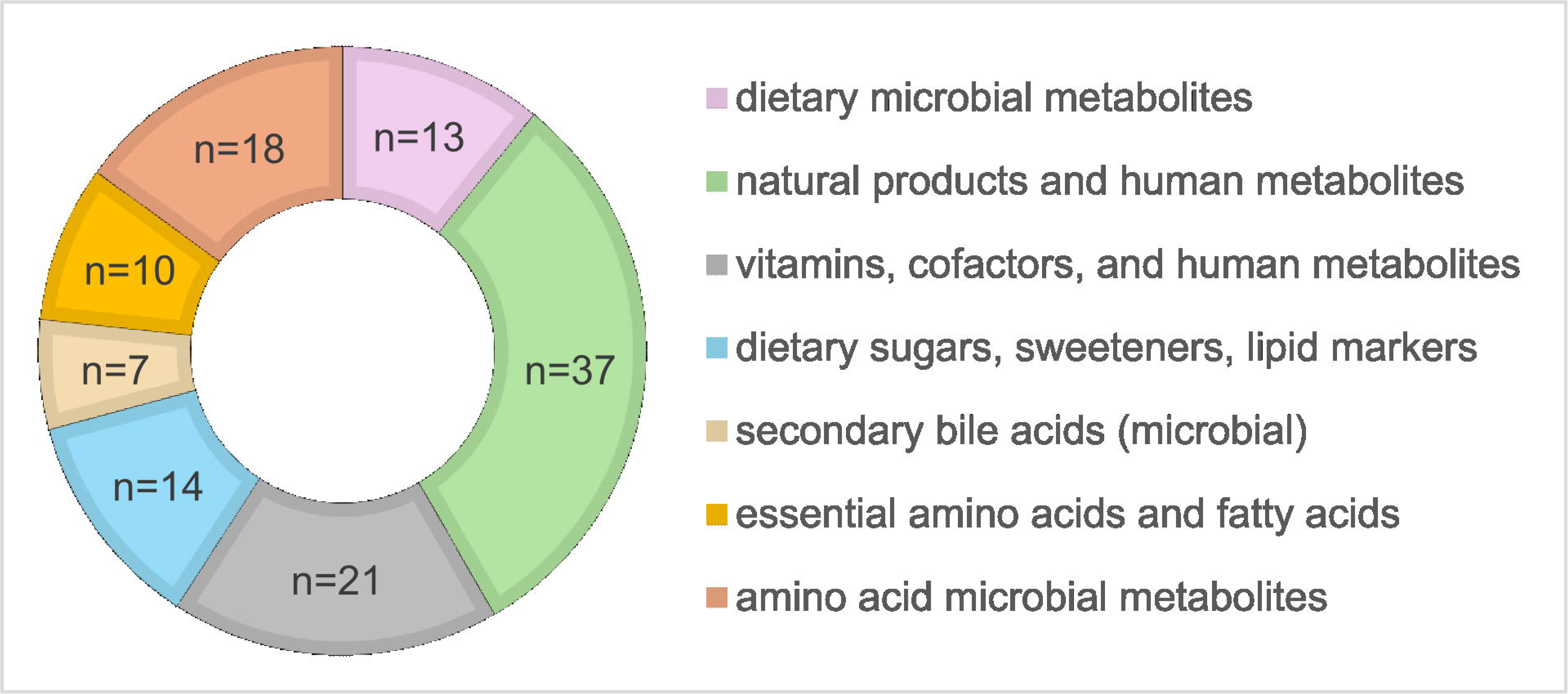
Class distribution of dietary and microbial compounds detected in the brain by the UCSD and Metabolon platforms. The dietary compounds included essential amino acids and fatty acids, as well as major metabolites of vitamins (e.g. pyridoxate from vitamin B6 products). The graph excluded endogenously produced sugars, amino acids, fatty acids, and downstream metabolites.

The DLPFC brain samples contained an array of compounds that originate in diet and retained their original form after their ingestion and absorption. These included the essential amino acids and fatty acids (detected in all brain samples) that could also be released from brain molecules that incorporated these compounds. Similarly, dietary carotenediols, and vitamins from the groups A, B, C, and E, were detected in the brain. The vitamins appeared either in their original form (from diet or microbial, such as vitamin Bs), or following human metabolism, especially that of vitamin B and C. Another micronutrient, queuine, found in 510 samples, is an essential modified nucleobase incorporated in tRNA, and is only produced by certain bacteria, hence it can originate in gut microbiota or the diet (plants, fungi, animals that salvaged it from bacteria). Additional natural products from plants and fungi were detected in the brain, such as piperine, solanidine, and the betaine-compounds ergothioneine, tryptophan-betaine (lenticin), and proline-betaine (stachydrine). A variety of dietary compounds underwent downstream metabolism by gut microbes and/or human biotransformation (phase I and II in the liver or other cells). The majority of these compounds were derived from aromatic amino acids and plant polyphenols, including several novel lignan metabolites detected by the UCSD platform. **Figure 5** illustrates potential dietary sources, and microbial and human biochemical transformations of 47 ingested compounds and metabolites detected in the brain.

**Figure 5.**
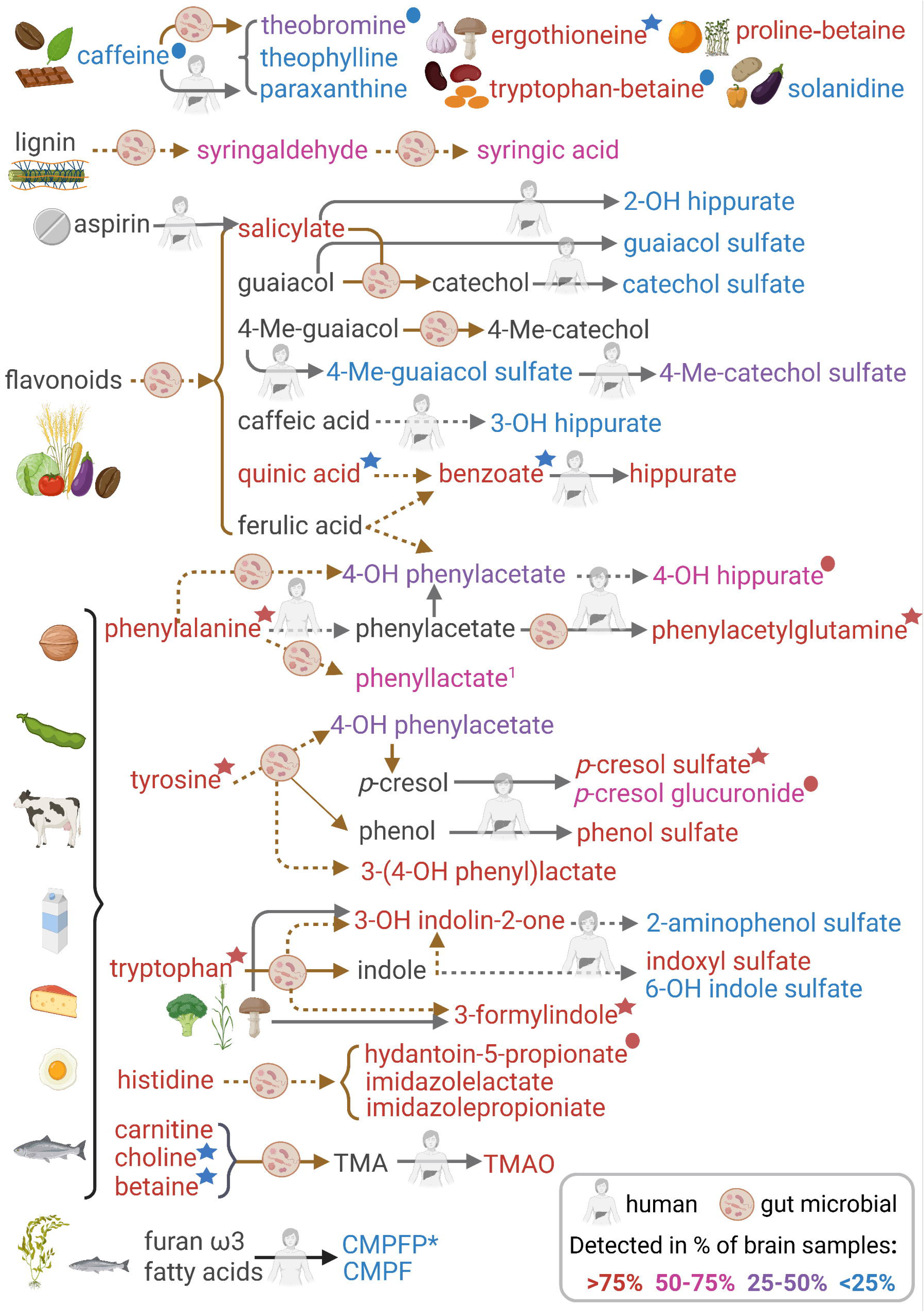
Dietary compounds and their suggested microbial and human metabolites detected in the brain. The text colour reflects the frequency of detection in the brain samples. Stars and bullets next to compound name indicate associations with cognition (significant or trend, respectively), with the colour indicating the directionality of correlation (blue – positive; red – negative). *Abbreviations:* OH, hydroxy; Me, Methyl; TMAO, trimethylamine N-oxide; CMPF, 3-carboxy-4-methyl-5-propyl-2-furanpropanoate; CMPFP*, 3-carboxy-4-methyl-5-pentyl-2-furanpropionate. *putative annotation of furan fatty acid metabolite. ^1^ Although not resolved by Metabolon, D-PLA is produced by microbes; L-PLA is produced endogenously. Created in BioRender.com.

Microbial digestion of plant fibre (lignin) produces syringaldehyde and syringic acid, both found in about two-thirds of the samples by the UCSD platform. Another class of microbial products detected in the brain was secondary bile acids, with almost half of the brain samples presenting with deoxycholic acid, and another six species (mostly conjugates) in less than 100 samples each. A slight overlap between dietary compounds and drugs was observed with salicylic acid (2-hydroxybenzoic acid), which can originate in plant phenolic compounds as well as from the analgesic aspirin (N-acetyl salicylate) and similar drugs. Also, the provitamin B5 dexpanthenol can be applied topically (also in skin products) or taken orally, especially via Total Parenteral Nutrition (TPN) as part of palliative care. A multitude of sugars were found in the brain, most of which probably originate in endogenous metabolism rather than consumption in food. However, a few outstanding examples are xylose (n=36), sucrose (n=500), and the artificial sweetener acesulfame found in about 10% of the samples. Some compounds can originate from several exposure sources. For example, 4-hydroxybenzaldehyde, found in plants (cereal), is a microbial metabolite of aromatic compounds including both natural benzenoids (e.g. mandelic acid) and industrial chemicals (BPA, toluene). It also utilised as an antimicrobial agent and is even a food chemical (flavouring agent). Other compounds are also used as food additives (benzoic acid, lactic acid, citric acid, taurine, maleic acid, fructose); however, when found in the brain and considering the dose contribution, they are more likely to originate in endogenous metabolism, plant, or microbial sources.

### 3.4. Environmental contaminants and industrial chemicals are possibly found in the brain

The two platforms together identified more than ten chemical compounds that possibly originate in environmental and lifestyle exposure (**Table S4**), although cross-contamination of the samples (false positives) cannot be completely ruled out. Some of those compounds retained their original industrial form, while others suggest metabolism by human or gut microbes. One of the most frequently detected compounds (>80% of samples) is 4-chlorobenzoic acid, potentially a microbial degradation product of chlorophenols such as the NSAID drug indomethacin, biphenyl industrial chemicals (PCBs), antiseptics, and even chlorinated pesticides. Over 20% of the samples showed methylparaben sulfate, a conjugated anti-microbial preservative utilised in PCPs, drugs, and food. Additional detected compounds with antimicrobial properties that can be used as PCP preservatives or in cleaning products include perillic acid, 2-Phenylethanol, and the benzyl quaternary ammonium compound Myristalkonium (also used as a preservative in eye drops). Other PCP ingredients suspected to be found in the brain are the emulsifier Cocamidopropylbetaine and the humectant PEG-4 (tetraethylene glycol). The antioxidant 2,4-DTBP (di-tertbutylphenol) is also employed as a preservative in PCPs and processed food, as well as to stabilise plastic. It was found by Metabolon in all brain samples, and although it is a common natural product from plants and microbes, it is frequently found in body fluids, potentially due to leaching from food packaging that incorporate tris(2,4-di-tert-butylphenyl)phosphite. Several plasticisers were also detected by the UCSD platform in brain samples, including bis(2-ethylhexyl) phthalate (DEHP) and dioctyl phthalate, both ubiquitous environmental contaminants. Other plasticisers / industrial solvents include Tris(1-chloro-2-propyl)phosphate, which is also a chlorinated flame retardant. The industrial chemicals and surfactants TMDD (2,5-dimethyl-3-Hexyne-2,5-diol) and PFOS (perfluorooctanesulfonic acid) were also found in the brain in more than 100 samples.

### 3.5. Applying BBB permeation in-silico models on exogenous compounds

To expand our understanding of the expected BBB permeation of the compounds annotated as possibly non-endogenous, we utilised LightBBB, an online prediction tool that receives the text chemical notation SMILES and uses a machine learning permeation model based on literature data for over 7000 compounds[43]. Of the over 130 potentially exogenous compounds, only 30 were predicted not to cross the BBB (see **Table S2-S4**), and many of those were human metabolites of exogenous compounds that can cross the BBB (such as ascorbate, pyridoxine, stachydrine, ibuprofen, paracetamol). Others could originate in local production rather than in diet, via nucleotide, energy and sugar metabolism, to mention a few. We further utilised a complementary *in silico* prediction tool of ADMET (Absorption, Distribution, Metabolism, Excretion, and Toxicity), AdmetSAR [44], integrated in DrugBank [45]. This model employs physico-chemical properties that dictate the potential of a compound to cross the BBB, including restricted passage of ions and polar solutes (>7 hydrogen bonds) and unconjugated macromolecules (MW>400 Da); passage of more lipophilic (lipid-soluble) small molecules (MW<400 Da), and very small hydrophilic ones (MW<150 Da). The ADMET results suggested a high likelihood of permeation (**Table S2-S4**) for some of the pharmaceuticals and natural products predicted as non-permeating by lightBBB (e.g., oxypurinol, warfarin, celecoxib, theophylline, carotenediols). The remaining non-permeating compounds included some key metabolites of high distribution in the DLPFC samples, which are further discussed.

In a reversed approach, the predicted permeability values may shed light on compounds missing in the brain and even be utilised to generate hypotheses related to brain health. For example, all seven methylated xanthine and uric acid metabolites derived from coffee, tea, and cacao natural products were not detected in the brain. They were all predicted as non-permeating, unlike their precursors caffeine, theobromine, theophylline, and paraxanthine (CYP2A1 product of caffeine, only in 16 brain samples). Supported by the low frequency of paraxanthine in brain, this also suggests little to no endogenous metabolism of the precursors with CYP2A1 and with xanthine dehydrogenase (XDH). XDH is an enzyme expressed in the DLPFC under normal conditions; however, it can be irreversibly converted to the enzyme xanthine oxidoreductase (XO) especially under elevated cellular calcium levels [46].

### 3.6. Brain exogenous compounds associated with cognition

In a previous publication[34] and by using the Metabolon platform and metabolomics data from DLPFC region in ROSMAP cohort, we have reported on widespread metabolic dysregulation associated with Alzheimer disease, spanning 298 metabolites from various AD-relevant pathways, mostly endogenous. These included alterations to bioenergetics, cholesterol metabolism, neuroinflammation, and metabolic consequences of neurotransmitter ratio imbalances. We have re-curated this dataset from Metabolon adding a list of recently identified compounds added to the Metabolon database and retested for correlations with cognitive function measured at last available assessment before death, utilising the complete dataset. For compounds that were found in at least 80% of the samples analysed by Metabolon, significant positive correlation with cognitive function was observed for several known antioxidants and health-supporting dietary compounds that also reflect the consumption of plant-based food and some animal-based food (see **Figure 5**).

These included ergothioneine, benzoic acid, quinic acid, betaine, carotenediol, vitamin C, and vitamins from the B group (pyridoxamine and riboflavin) and E groups (tocopherols). In contrast, a significant negative correlation with cognition was found for large neutral amino acids, indole and cresol microbial products (3-formylindole, p-cresol sulfate), as well as metabolites of vitamin C (2-O-methylascorbic acid, dehydroascorbate, threonate), and the sugars erythritol and sucrose. This confirms earlier findings.

We next conducted a complementary analysis using less stringent missingness criteria to include compounds detected in fewer than 80% of samples. The analysis also suggested that additional compounds, including tryptophan-betaine and retinol (vitamin A), showed a positive trend with better cognition. Furthermore, several compounds were associated with lower cognition, including the microbial products p-cresol glucuronide conjugate, hydantoin-5-propionate, 4-hydroxyhippurate, and the putative secondary bile acid glycolithocholate sulfate (**Table S5**). These findings should be interpreted as exploratory until confirmed in additional ROS/MAP brains and independent cohorts.

## 4. DISCUSSION

### 4.1 BBB permeation and link to DLPFC metabolites

The entry of chemicals into the brain is regulated to maintain homeostasis and enable the supply of nutrients and other solutes from the blood. Passive diffusion through the BBB is ruled by physico-chemical properties, while active passage utilises dedicated carrier-mediated transporters such as the monocarboxylic acid transporter (MCT-1), Large-Neutral amino acid (LAT-1), basic amino acid (CAT-1), glucose transporter (GLUT1), among others [47, 48]. Some drugs that target the CNS draw on chemical similarity. For example, the anticonvulsant drug gabapentin (found in 43 brain samples), crosses via LAT1 due to a chemical structure (cyclic gamma-amino acid) that mimics alpha-amino acids. Affinity to transporters can also affect the rate of active brain efflux vs. influx, contributing to the net presence of chemicals in the brain.

Our results employing the lightBBB and ADMET permeation prediction left a few interesting compounds and direct precursors as non-permeating, and literature indicates specific carriers for some (see **Tables S2-S3**). Queuine, the essential modified nucleobase obtained via diet and (mainly) gut microbiota, reaches the brain via a specific solute carrier (SLC35F2) [49]. It is specifically enriched in the brain as a substrate for the enzyme guanine-queuine tRNA transglycosylase [50]. Examining some of the metabolites that were associated with better cognitive function in our cohort, benzoic acid may pass through the BBB via the pH-dependent monocarboxylic acid transporter (MCT1 SLC16A1) [51], and other small phenolic compounds may follow suit (e.g. salicylic acid). In comparison, ergothioneine is suggested to cross via an almost-exclusive organic cation transporter (OCTN1 SLC22A4) [52, 53]. Of the compounds that were negatively associated with cognition, three metabolites of vitamin C are not predicted to cross the BBB, unlike their precursor. This may suggest brain endogenous metabolism of vitamin C, rather than transfer of the metabolites. The disaccharide sucrose is also too hydrophilic for passive diffusion and has no known active carriers. This raises the possibility of BBB endothelial dysfunction allowing “leakage” of unexpected compounds into the brain [54]. Sucrose is considered a marker of BBB permeability, and its finding in 97% of samples suggests not only an association with the degree of BBB endothelial dysfunction [55], but also increased intestinal epithelial permeability (“leaky gut”) [56] (this disaccharide is expected to be digested in the small intestines into monosaccharides, with leftovers fermented by gut microbiota or excreted).

The dietary sugar alcohol erythritol (in all brain samples) is easily absorbed in the small intestines, is polar with no dedicated BBB carrier, and only minor endogenous production via the pentose-phosphate pathway. An *in-vitro* study linked it to microvascular endothelial damage [57], however, rather than increased BBB permeation, it could suggest increased endogenous production due to metabolic stress [58].

This recent debate on biochemical interpretation highlights the continuous progress in the research of chemical delivery to the brain, which requires further evidence *in-vitro* and *in-vivo*, alongside *in-silico* models. Moreover, increased BBB permeability due to health state suggests a bias in interpretation of the presence of brain metabolites, possibly reflecting merely an indirect health effect rather than disease biochemistry. While BBB dysfunction increases due to aging, inflammation, and disease, the endothelial membrane integrity can also be disrupted by compounds such as bacterial lipopolysaccharides, heavy metals, and surface-active chemicals (such as microbial secondary bile acids) [59]. Our finding of twelve primary and secondary bile acids in the brain samples demonstrates their ability to cross the BBB, owing to their physicochemical properties and surfactant activity (possibly resulting in membrane disruption). The microbial-human co-metabolite TMAO (in all brain samples) and other metabolites can induce downregulation of tight junction proteins [60] that support the integrity of the endothelial bilayer [54]. BBB dysfunction can also be temporarily induced using medical stimuli to increase CNS drug delivery to the brain [61], including “relaxing” the regulation of tight junction via osmotic or chemical agents (e.g., histamine) [48].

All the annotated exposure chemicals detected in the brain (**Table S4**) could potentially cross the BBB according to the lightBBB model results. Nevertheless, some have not been discussed as accumulating in the brain until very recently. The persistent environmental pollutant PFOS used in a wide range of domestic, industrial, and laboratory products [62] was found in 112 brain samples, aligning with previous reports of detection in the brain [63], and possible association of the class PFAS with cognitive disturbances [64–66].

Methylparaben (its sulfate-conjugate was detected in more than 20% of the samples), is considered safe as a food preservative up to a certain concentration. Evidence for its brain accumulation is only recent [67], after years of assuming that it is hydrolysed and rapidly excreted in urine [68]. Specifically, it was found in higher concentrations in the hypothalamus and increased in people with obesity [67]. Phthalates, especially DEHP detected in >60% of samples, is a contaminant of high interest. Rat exposure to DEHP during gestation and lactation was associated with increased Tau phosphorylation and cognitive dysfunction at older age [69]. In the NHANES cohort, DEHP was the leading phthalate associated with insulin resistance [70].

### 4.2 Alternative routes into the brain

The presented discussion assumes that the route to the brain was through crossing the BBB; however, there are additional routes to the brain that complicate the study of body-brain metabolic links. The intranasal introduction of chemicals can bypass the BBB, if not eliminated in their course or they enter the blood circulation via lungs or GI tract. More importantly, there is a flow of compounds through the blood-CSF endothelial layer and then possibly from the CSF to the brain via CSF-perivascular pathways [71]. A potential pathway is via the CNS meningeal lymphatic vessels that reach the brain and support the CNS homeostasis. This system was recently characterised to work in tandem with the glymphatic route to enable the para-vascular exchange of molecules between the CSF and the interstitial fluid [72]. Although it was described in the context of facilitating the clearance of molecules out of the brain, it may contribute to transport from the lymph system and the CSF into the brain.

### 4.3 DLPFC metabolites linked to brain health and cognition

The exogenous brain compounds that were associated with cognition in the cohort exhibit a variety of bioactivities, and here we concentrate on selected few. One of the strongest associations with better cognition was seen with ergothioneine (thiolhistidine-betaine), a bacterial and fungal product [73] found at high levels in dietary mushrooms [74], beans, and oats [75]. It is considered an antioxidant with neuroprotective properties [59, 76] and in the Rotterdam study it emerged as the lead blood biomarker associated with better cognition [77]. Blood ergothioneine decreased in people with frailty and cognitive decline [78] and was trialled as a supplement in people with MCI [79]. Although its exact mechanism is under investigation, *in vitro* studies demonstrated interactions with signalling cascades related to aging and oxidation, and promotion or neuronal stem differentiation through activation of several signalling pathways [80]. Quinic acid, a cyclic polyol from coffee and other plants, also exhibited antioxidative, anti-microbial, and anti-neuroinflammatory activities [81, 82], and supported neuronal cell proliferation *in-vitro* [83].

Carotenediols can be found in different food sources and have antioxidative activity; however, their BBB permeation is partial. In The TILDA study, plasma carotenediol was positively associated with better cognition in >4000 people aged >50 years [84]. Carotenoid supplementation was widely reviewed for alleviating AD symptoms [85]. Cognitive decline and AD were associated not only with higher brain levels of p-cresol sulfate but also higher plasma levels [86–88]. This gut microbial metabolite of tyrosine (and possibly benzenoids as well) is considered neurotoxic due to NMDA receptor alteration [89], and inhibition of dopamine–beta-hydroxylase which converts dopamine to norephinephrine (with ascorbic acid and copper as coenzymes) [90, 91]. p-cresol was also linked to defective myelination, increased dopamine turnover, and decreased differentiation to oligodendrocytes [92]. Lastly, the association of ascorbic acid (vitamin C) with better cognition and its derivatives with worse cognition possibly reflects the involvement of vitamin C in critical neurotransmission reactions as the example above, as well as neurogenesis, and it was the target of trials, studies, and reviews [93].

### 4.4 Study Limitations

The chemical profile of the DLPFC brain region may differ from other brain regions and presents only a part of the brain metabolome atlas. The lipophilic nature of brain tissue makes it highly susceptible to trapping contaminants from plastic containers, etc. Therefore, the presence of some industrial chemicals in the brain could be due to exposure during post-mortem handling, sample preparation, and analysis in each chemical lab. Despite the careful protocols applied in this study, false positive results that identify chemicals due to cross-contamination may not be fully mitigated by the appropriate analytical approaches (pre- and post-analysis). Nevertheless, we reported the findings of interest, acknowledging that they could benefit from confirmation in independent cohorts and by complementary methods.

### 4.5 Conclusion

The brain metabolic profile presented here highlights the presence of over 200 exposome-related compounds from the environment, drugs, diet, and gut microbial metabolites. The presence of exposome-related compounds in human brains at a physiologically relevant concentration can have profound effects on brain function and may influence brain health. A detailed mapping of the metabolome in different brain regions and its linking to peripheral metabolism across the trajectory of disease, can provide great insights into peripheral central links in AD pathogenesis. Exploration of longitudinal studies accompanied by comprehensive lifestyle metadata can reveal the extent of these lifetime exposures. While the correlation between blood and brain metabolome is still under investigation, all evidence points to the importance of connecting body and brain in the study of CNS diseases. Expanding this research field will provide totally new insights about disease mechanisms, and opportunities for drug interventions where modifiable lifestyle factors are used in combination therapy approaches.

## Data Availability

The results
published here are in whole or in part based on data obtained from the AD Knowledge Portal (https://adknowledgeportal.org)

## ACKNOWLEDGEMENTS

The Alzheimers Gut Microbiome Project (AGMP) and the Alzheimer’s Disease Metabolomics For up-to-date information on the partners, visit https://fnih.org/our-programs/accelerating-medicines-partnership-amp/amp-alzheimers-disease-2-0/). As such, the investigators within the AGMP and the ADMC, not listed specifically in this publication’s author’s list, provided analysis-ready data, but did not participate in designing the study, conducting the analyses or writing this manuscript. A listing of AGMP Investigators can be found at https://alzheimergut.org/meet-the-team/. A listing of ADMC investigators can be found at: https://sites.duke.edu/adnimetab/team/.

We acknowledge the editorial support of Jon Kilner, MS, MA (Pittsburgh).

## Abbreviations

AD: Alzheimer’s Disease
ADMC: Alzheimer’s Disease Metabolomics Consortium
ADNI: Alzheimer’s Disease Neuroimaging Initiative
ADMET: Absorption, Distribution, Metabolism, Excretion, and Toxicity
AGMP: The Alzheimers Gut Microbiome Project
BA: Bile Acids
BBB: Blood Brain Barrier
CNS: Central Nervous System
DEHP: bis(2-ethylhexyl) phthalate
DLPFC: Dorsolateral Prefrontal Cortex
GNPS: Global Natural Product Social Molecular Networking
MAP: Rush Memory and Aging Project
MCI: Mild Cognitive Impairment
MIND: Mediterranean-DASH dietary intervention
PCP: Personal Care Products
ROS: Religious Orders Study
ULPC: Ultra-Performance Liquid Chromatography
XDH: Xanthine Dehydrogenase

