## Supplementary Methods for "Exposome contribution to the brain metabolome: importance of body brain connection"

**Exposome contribution to the brain metabolome: body brain connections can impact brain health**

### Supplementary Methods

#### 1. Collection and preparation of brain samples

##### 1.1. Procedure at the time of autopsy

The hemisphere selected for freezing was sliced into 1cm coronal slabs, photographed, and frozen between two brass plates which had been pre-cooled to -80°C then laid out on a bed of dry ice. Once frozen, each slab was stored in individual, recloseable plastic bags labelled with the case number and participant ID. The bags were placed in 3” fibreboard boxes and stored at -80°C for future use.

##### 1.2. Procedure at the time of collection for distribution

At the time of collection, the technician prepared a bed of dry ice on a cart, located the first ID in Freezerworks, found the position, and pulled the sample. The brain slabs, in individually labelled bags, were laid out on a bed of dry ice. The technician checked the project ID on the sample and on the cryovial label against the spreadsheet and Freezerworks. Once they were confirmed to match, he or she placed the label on the cryovial, set the vial on a scale, and tared the scale. The technician then identified the region of interest using a Brodmann Area map or brain atlas and used one of two instruments to perform the dissection: The fine-toothed razer saw (for cortical regions) or the jewelers saw with diamond wire (for subcortical structures). Once the dissection was complete, the piece of tissue was placed in a 2mL sterile polypropylene cryovial (Heathrow Scientific, IL, USA). The vial weight was documented, and the sample to be distributed was filed in a labelled box sitting on dry ice. In Freezerworks a sub-aliquot was created, assigned a position, and flagged for distribution. The brain was then packed back up. The technician changed gloves and returned the brain to its designated position in the freezer. All instruments and cutting surfaces were cleaned with 70% ethanol, and the process started over with the next ID on the list. No reagents or preservatives were used at any time when handling fresh-frozen tissue.

#### 2. Metabolomics analysis - Metabolon

##### 2.1. Sample Extraction

Upon receipt, samples were immediately stored at -80°C until processing. Samples were prepared using the automated MicroLab STAR® system from the Hamilton Company. Several recovery standards were added prior to the first step in the extraction process for quality control (QC) purposes. To remove protein, dissociate small molecules bound to protein or trapped in the precipitated protein matrix, and to recover chemically diverse metabolites, proteins were precipitated with methanol under vigorous shaking for 2 min (Glen Mills GenoGrinder 2000), followed by centrifugation. The resulting extract was divided into several fractions: two for analysis by two separate reverse phase (RP)/ultra-performance liquid chromatography (UPLC(-MS/MS methods with positive ion mode electrospray ionization (ESI), one for analysis by RP/UPLC-MS/MS with negative ion mode ESI, and one for analysis by hydrophilic interaction chromatography (HILIC)/UPLC-MS/MS with negative ion mode ESI. Samples were placed briefly on a TurboVap® (Zymark) to remove the organic solvent. The sample extracts were stored overnight under nitrogen before preparation for analysis.

2.2. Instrumental Analysis

All methods utilized a Waters ACQUITY UPLC and a Thermo Scientific Q-Exactive high resolution/accurate mass spectrometer interfaced with a heated electrospray ionization (HESI-II) source and Orbitrap mass analyzer operated at 35,000 mass resolution. The sample extract was dried then reconstituted in solvents compatible to each of the four methods. Each reconstitution solvent contained a series of standards at fixed concentrations to ensure injection and chromatographic consistency. One aliquot was analyzed using acidic positive ion conditions, chromatographically optimized for more hydrophilic compounds. In this method, the extract was gradient eluted from a C18 column (Waters UPLC BEH C18-2.1x100 mm, 1.7 µm) using water and methanol, containing 0.05% perfluoropentanoic acid (PFPA) and 0.1% formic acid (FA). Another aliquot was also analyzed using acidic positive ion conditions; however, it was chromatographically optimized for more hydrophobic compounds. In this method, the extract was gradient eluted from the same aforementioned C18 column using methanol, acetonitrile, water, containing 0.05% PFPA and 0.01% FA, and was operated at an overall higher organic content. Another aliquot was analyzed using basic negative ion optimized conditions using a separate dedicated C18 column. The basic extracts were gradient eluted from the column using methanol and water with 6.5mM Ammonium Bicarbonate at pH 8. The fourth aliquot was analyzed via negative ionization following elution from a HILIC column (Waters UPLC BEH Amide 2.1x150 mm, 1.7 µm) using a gradient consisting of water and acetonitrile with 10mM Ammonium Formate, pH 10.8. The MS analysis alternated between MS and data-dependent $MS^n$ scans using dynamic exclusion. The scan range varied between methods but covered 70-1000 mass to change ratio (*m/z*).

2.3. Feature Extraction, Data Processing and Annotation

Raw data was extracted, peak-identified and QC processed using Metabolon's hardware and software. Peaks were quantified using area-under-the-curve. For studies spanning multiple days, a data normalization step was performed to correct variation resulting from instrument inter-day tuning differences. Compounds were identified by comparison to library entries of purified standards or recurrent unknown entities, incorporating retention time/index (RI), mass to charge ratio (*m/z)*, and chromatographic data (including MS/MS spectral data). Library matches for each compound were checked for each sample and corrected if necessary.

2.4. Statistical analysis

Initial statistical analyses of the DLPFC Metabolon dataset in the ROSMAP cohort were reported previously [1]. For the present study, after re-curation of the dataset to incorporate newly annotated compounds from the Metabolon database, associations with cognitive function were re-evaluated using two complementary analytical strategies.

In the primary analysis, only metabolites detected in at least 80% of samples were included. Probabilistic quotient normalization was applied to correct for sample-wise variation, followed by log2 transformation. Missing values were imputed using k-nearest neighbors (kNN). Sample-level outliers were identified and removed using the local outlier factor (LOF) method across a range of neighborhood sizes (k = 5, 10, 20, 30, 40, 50). Metabolite-level outliers were detected using a quantile-based threshold (2.5th/97.5th percentiles), set to missing, and re-imputed with kNN. The effects of concomitant medications were regressed out using LASSO regression with 5-fold cross-validation, selecting the penalty at lambda.min; Alzheimer's disease and neurological drug classes were excluded from this correction to avoid removing variance of interest. Association analyses were conducted using linear models adjusted for APOE genotype, sex, age at death, education, postmortem interval, and body mass index.

In a complementary analysis designed to facilitate the evaluation of metabolites with higher missingness, probabilistic quotient normalization was again applied, followed by log2 transformation. Missing values were imputed by substituting the minimum observed value for each metabolite. The effects of concomitant medications were regressed out using LASSO regression with 5-fold cross-validation, selecting the penalty at lambda.min; Alzheimer's disease and neurological drug classes were excluded from the correction to avoid removing variance of interest. Association analyses were conducted using the same linear models and confounders as for blood metabolomics. In both analyses, p-values were corrected for multiple testing using the Benjamini–Hochberg false discovery rate.

### 3. Metabolomics analysis - UCSD

3.1. Sample Extraction.

Brain samples were plated onto wooden swabs and stored at −80°C. For extraction, each swab was transferred into a sterile Qiagen tube and supplemented with 400 µL LC-MS grade water. Samples were mechanically homogenized using a TissueLyser (25 Hz, 5 min) to release tissue material from the swabs, followed by centrifugation at 16,000 rpm for 5 min. Swabs were then removed from the tubes. To extract organic-soluble metabolites, 400 µL of 100% methanol containing 1 µM sulfamethazine (internal standard) was added to the remaining tissue lysate pellet. Samples were centrifuged again at 16,000 rpm for 5 min, and supernatants were incubated at -20°C for 30 min to enhance protein precipitation. A 200 µL aliquot of the supernatant was transferred into a shallow-well collection plate and dried to completeness on a vacuum concentrator. Dried extracts were sealed and stored at -80°C until resuspension in 150 µL of 50:50 acetonitrile/water for LC-MS analysis.

#### 3.2. Instrumental Analysis

Extracts (5 µL) were analyzed using a Vanquish UHPLC system coupled to a Q Exactive quadrupole-orbitrap mass spectrometer (Thermo Fisher Scientific). Chromatographic separation was performed on a Kinetex C18 column (50 × 2.1 mm, 1.7 µm, 100 Å; Phenomenex) equipped with a SecurityGuard C18 cartridge (2.1 mm ID) maintained at 30°C. Mobile phases were 0.1% formic acid in water (A) and 0.1% formic acid in acetonitrile (B) with the following gradient (0.5 mL/min): 0-1.0 min, 5% B; 1.0-7.0 min, 5-100% B; 7.0-7.5 min, 100% B; 7.5-8.0 min, 100-5% B; 8.0-10.0 min, 5% B. The mass spectrometer was operated with positive heated electrospray ionization with the following settings: sheath gas, 53 AU; auxiliary gas, 14 AU; sweep gas, 3 AU; auxiliary gas temperature, 400 °C; spray voltage, 3.5 kV; capillary temperature, 269 °C; S-lens RF level, 50 V. Full scan MS1 spectra were acquired from *m/z* 100-1500 at 35,000 resolution (at *m/z* 200), with a maximum injection time of 100 ms and AGC target of 5E5. Data-dependent MS/MS was performed on up to five precursors per MS1 scan using a 3 *m/z* isolation window, 0.5 *m/z* isolation offset, stepped normalized collision energies (20/30/40%), 35,000 resolution, 100 ms maximum injection time, AGC target of 5E5, minimum AGC of 5E3, apex trigger of 2-15 s, and dynamic exclusion of 10 s.

#### 3.3. Feature Extraction and Data Processing

Raw files were converted to mzML format using MSConvert (ProteoWizard) and processed in MZmine 2. Mass detection employed noise thresholds of 5E4 for MS1 and 1E4 for MS2. Chromatogram construction used a mass tolerance of 0.001 *m/z* or 10 ppm, a minimum of five consecutive scans, and a minimum peak height of 1.5E5. Chromatograms were deconvoluted using the local-minimum search algorithm (search range 0.2 min; minimum peak-top/edge ratio 1.5; maximum peak duration 1.5 min). Isotopic peak grouping applied tolerances of 5 ppm (*m/z*) and 0.15 min (retention time). Features were aligned using 0.001 *m/z* or 10 ppm mass tolerance and 0.3 min retention-time tolerance, followed by gap-filling with 20 ppm *m/z* and 0.1 min retention-time tolerances. The resulting feature table (.csv) and MS/MS spectra (.mgf) were exported without further post-processing. Blank subtraction was performed by removing features whose mean peak areas in brain samples were less than three times the mean peak areas observed in extraction blanks. Additionally, any feature with MS/MS spectra detected in blank swabs was removed. Detection frequency was then calculated for each feature as the percentage of samples with peak area higher than 1E4. Feature annotations were assigned using the GNPS spectral library [2] (job link: https://gnps2.org/status?task=c3382680e42b44f9bb2daf1086139ba3).

3.4. Reference Data–Based Source Annotation

To contextualize metabolite origins, spectra were queried against curated reference datasets containing >60,000 microbial monocultures, ~3,500 food extracts, and ~500 personal-care products using the Fast Search API within the microbeMASST framework (https://github.com/robinschmid/microbe_masst) [3, 4]. Searches used a 0.05 Da mass tolerance, minimum cosine of 0.7, and at least five matched peaks. A feature was assigned a potential source if fewer than 5% of its spectral matches originated from blanks or QC samples within the curated reference datasets for that source. Drug and drug-metabolite annotations were derived using the curated GNPS Drug Library[5]. Features matching uniquely to a single source domain were labelled accordingly; features matching multiple domains were labelled "multiple."

3.5. Body-Part Distribution Across Public Metabolomics Datasets

To evaluate whether brain-detected metabolites also appear in other human tissues, ROSMAP MS/MS spectra were searched across all public untargeted metabolomics datasets in GNPS/MassIVE [6], MetaboLights, and the Metabolomics Workbench via the Fast Search API. Matches were merged with ReDU ("Reanalysis of Data User Interface"), which provides harmonized sample metadata. Only matches to human datasets ("9606 | Homo sapiens" in the NCBITaxonomy field) with annotated body-part information ("UBERONBodyPartName" in ReDU) [6] were retained. An UpSet plot summarizing the co-occurrence of features across body-part combinations was generated using the ComplexUpset R package (v1.3.3).
